# Estimation of fine-scale spatial heterogeneity in treatment effects in a large cluster-randomized trial

**DOI:** 10.64898/2026.11.03.26367216

**Authors:** Ariktha Srivathsan, Mia A. Navarro, Ahmed M. Arzika, Amza Abdou, Ramatou Maliki, Almou Ibrahim, Diallo Beidi, Nasser Galo, Nasser Harouna, Alio M. Karamba, Moustapha Abarchi, Brittany Peterson, Pearl Anne Ante-Testard, Elodie Lebas, Travis C. Porco, Marc Lipsitch, Kieran S. O’Brien, Thomas M. Lietman, Benjamin F. Arnold

## Abstract

Treatment effects in randomized trials may vary geographically, and estimating this variation is a broadly generalizable problem for large-scale, cluster-randomized trials. We conducted a secondary analysis of AVENIR, a cluster-randomized, placebo-controlled trial of biannual azithromycin distribution among children aged 1–59 months in Dosso, Niger (2020–2023). We assessed whether the survival benefit of mass azithromycin distribution varied geographically using Poisson models to estimate spatially continuous child mortality and average treatment effects (ATE). Among 1390 villages (276,452 person-years; 3635 deaths), the overall mortality rate was 13.1 and the ATE was −2.6 deaths per 1000 person-years. Mortality was highest in the south, whereas treatment benefit was concentrated in the southeast, following a northwest–southeast gradient. Aggregation of data to 160 primary health catchment areas preserved broad spatial patterns in mortality but attenuated local heterogeneity, suggesting that health catchment monitoring is sufficient for broad mortality surveillance. Examining unit-level, spatially predicted treatment effects as a function of mortality rate demonstrated that underlying mortality captured much, but not all, of the heterogeneity in treatment effects. The analysis demonstrates how to estimate localized treatment effects in large cluster-randomized trials and provides new insights into spatial heterogeneity in mortality and benefits of azithromycin distribution in Niger.

## Introduction

A treatment’s effect might vary across a trial’s study area: geographic location integrates differences in ecology, access to care, and population composition that can plausibly modify an intervention’s benefit^1^. Estimating where, and by how much, treatment effect varies geographically is therefore a broadly generalizable problem for large-scale randomized trials conducted across dispersed communities. Child survival interventions in high-mortality settings provide a motivating example due to their large scale and having plausible geographic variation in effects. Geographic inequities in child survival persist even as under-5 mortality rates have declined globally.^2^ Analyses of population-based data show that child mortality is heterogenous at subnational scales, with high-risk areas often clustering spatially rather than distributing uniformly across countries or regions.^3–7^ In the Sahel and nearby West African settings, this geographic patterning is plausibly shaped by ecological and livelihood conditions, including climate variability, vegetation, malaria ecology, population density, and access to care, all of which may influence child survival and intervention effects through nutrition, infection, and service-access pathways.^8–11^

In high-mortality settings, randomized controlled trials have shown that twice-annual mass drug administration (MDA) of azithromycin to children aged 1–59 months reduces all-cause mortality.^12,13^ AVENIR (*Azithromycine pour la Vie des Enfants au Niger: Implementation et Recherche*), a cluster-randomized trial in the Dosso and Tahoua regions of Niger, found that biannual distributions to children 1–59 months reduced mortality by 14% relative to placebo, while distributions limited to infants 1–11 months did not achieve a statistically significant reduction.^14^ Subgroup analyses from AVENIR and related trials of azithromycin MDA in Africa have examined whether treatment effects vary by baseline mortality rate^15^, nutritional status^16–18^, and access to primary care.^19^ Pooled analyses across trials in Ethiopia, Malawi, Niger, and Tanzania could not rule out modest effect modification of MDA by underlying mortality rate,^20,21^ suggesting that the mortality benefit of azithromycin MDA may vary across populations and settings. However, the analyses have not examined fine-scale spatial heterogeneity in treatment effects within a region.

The literature on heterogeneous treatment effects in randomized trials has focused predominantly on clinical trials and patient-level covariates rather than geographic structure as a source of variation.^22^ When geography is considered as an effect modifier in trials, it is often assessed at a high spatial scale, such as country or region.^23,24^ Spatial modeling in cluster randomized trials has focused on estimation of heterogeneous treatment effects within and between individual villages, at very small spatial scale, with an emphasis on between-cluster spillover effects.^25–30^ Geospatial models are widely used in disease risk mapping,^31–34^ but modeling continuous treatment effect surfaces at district- or regional scales has, to the best of our knowledge, only been well-defined within geographically pair-matched trial designs.^35^

High-mortality countries are now implementing national-scale azithromycin programs for child survival, and ongoing treatment-effect estimation is not feasible once MDA is universal. If effects vary within a region, understanding the relevant spatial scale of effect heterogeneity and spatial resolution of mortality measurement required to detect it could help inform future surveillance and decision making. For example, embedding childhood mortality monitoring within existing primary healthcare catchment areas may offer a pragmatic way to track mortality as programs expand, provided that catchment-level aggregation retains enough spatial resolution.

Here, we extend spatially resolved treatment-effect estimation to AVENIR, asking whether and where geography modifies the mortality benefit of azithromycin across Niger’s Dosso region. We assess whether spatial aggregation of mortality measurement to 160 healthcare catchment areas preserves the spatial patterns in mortality detectable when analyzed at the village level, with implications for programmatic surveillance. As an additional contribution, we assess to what extent a location’s mortality rate serves as a proxy for its anticipated benefit in terms of child survival, because, unlike treatment effects, mortality rates can be measured outside the context of a randomized trial.

## Methods

This is a secondary analysis of AVENIR^36^ (NCT04224987), an adaptive, cluster-randomized, placebo-controlled trial conducted in Niger to evaluate the effect of MDA with azithromycin on child mortality.

### Trial oversight

The trial received ethical approval from the Comité National Éthique pour la Recherche en Santé in Niger and the University of California, San Francisco Institutional Review Board. Community leaders gave verbal informed consent before study activities began, and verbal consent was also obtained from heads of household for census activities. Caregivers of eligible children gave verbal informed consent for treatment administration.

### Trial design and participants

Rural and peri-urban villages in the Dosso region of Niger with populations between 250 and 2,499 people were eligible for inclusion in the study. Eligible villages were randomized to one of three arms: a child arm (azithromycin administered to children aged 1–59 months), an infant arm (azithromycin administered to children aged 1–11 months and placebo to children aged 12–59 months), and a placebo arm (placebo administered to children aged 1–59 months). Because the infant arm showed no statistically significant effect on mortality in 1–59 month-old children,^14^ villages randomized to the infant and placebo arms were combined into a single control group for the present analysis, and the child arm constituted the treated group. Additionally, only villages from the trial’s first randomization that completed 24 months of follow-up were included in the present analysis, all located in the Dosso region (**Figure 1**). This pooling and restriction were done to maximize available person-time and support fine-scale spatial estimation of mortality, a rare outcome.

**Figure 1:**
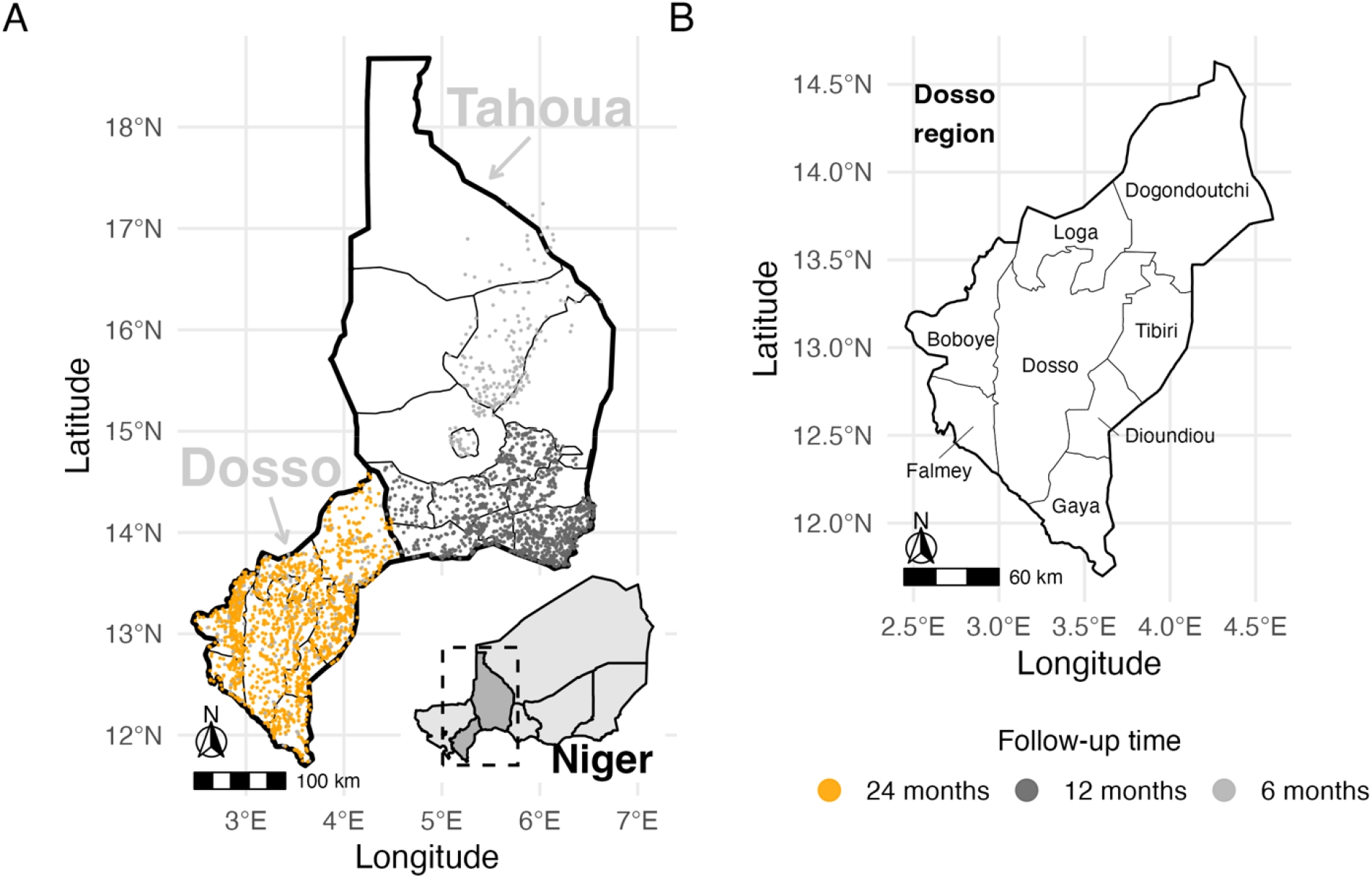
AVENIR (Azithromycine pour la Vie des Enfants au Niger: Implementation et Recherche) study area in Niger, 2020-2023. (A) The trial area includes the Dosso and Tahoua regions of Niger. The country of Niger is shown in the inset, with included regions shaded in dark grey. Points represent rural and peri-urban villages that were included in the trial (N = 2920). Of these, yellow points (N = 1390) represent villages that had 24 months of follow-up and hence included in this spatial analysis. (B) All the villages included in this analysis are in the Dosso region of Niger. This map shows the 8 departments within the Dosso region: Boboye, Dioundiou, Dogondoutchi, Dosso, Falmey, Gaya, Loga, and Tibiri. Administrative boundaries used in maps were sourced from the United Nations Office for the Coordination of Humanitarian Affairs (OCHA) *Common Operational Datasets – Administrative Boundaries*, accessed via the Humanitarian Data Exchange on 10 March 2026, and are licensed under CC BY-IGO 3.0 (https://data.humdata.org/dataset/cod-ab-ner). Alt text: Maps of the trial study area in Niger, showing village locations by follow-up duration in the Dosso and Tahoua regions and the eight departments of the Dosso region.

The outcome of interest was the child mortality rate, defined as deaths per 1,000 person-years among children aged 1–59 months, ascertained through census activities undertaken every six months. The average treatment effect (ATE) was defined as the incidence rate difference (treatment minus control), such that negative values indicate a beneficial effect of azithromycin.

Department (district) level mortality rates were estimated separately for treatment and control villages. To assess heterogeneity in child mortality rates across the eight departments in the Dosso region, a Poisson generalized linear model with a log link was fit with department and treatment group as fixed effects and the log of person-years as an offset, that is, log(µ_i_) = log(PT_i_) + β_o_ + β’ dept_i_ + β_1_Trt_i_, where μ□ is the expected death count in village i, PT□is person-years at risk, dept_i_ represents the fixed effect for the department of village i, and *Trt*_*i*_ is an indicator variable for treatment group of village i. A likelihood ratio test (LRT) compared this model against a Poisson model with treatment indicator and intercept only, and the same offset, log(µ_i_) = log(PT_i_) + β_o_ + β_1_Trt_i_, with the test statistic referred to a χ^2^ distribution on seven degrees of freedom. The intraclass correlation coefficient (ICC) quantifying the clustering of child mortality within departments was estimated from a Poisson generalized linear model with a fixed effect for treatment, a random intercept for department 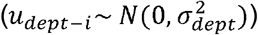, and the log of person-years as an offset, log(µ_i_) = log(PT_i_) + β_o_ + u_dept− i_ + β_1_Trt_i_ ; 95% confidence intervals were obtained from 1000 bootstrap draws.

### Geospatial models

Spatial variation in child mortality and the ATE was characterized using four Poisson generalized linear mixed models (GLMMs) with Matérn-structured spatial random effects and an offset for the log of person-years of follow-up, fit using maximum likelihood. The Matérn covariance function was used because it defines correlation as a continuous function of distance, which is more natural for irregularly spaced point locations than neighborhood-based alternatives, consistent with recommendations for spatially continuous trial data.^37^ The covariance is governed by two parameters: a spatial range, which sets the distance over which correlation between villages decays, and a smoothness parameter (*v*), which sets how smoothly the effect varies across space.

Four GLMMs were built sequentially, each relaxing an assumption of the preceding specification (**Supplementary table 2**). The first model (baseline spatial model) included an intercept, a binary treatment group indicator, and a Matérn spatial random effect, encoding the assumption that baseline mortality risk varies smoothly across space but that the treatment effect is spatially constant. This model can be written as

log(µ_i_) = log(PT_i_) + β_o_ + β_1_Trt_i_ + *w*(s_*i*_), where *w*(*s*_*i*_) is a mean-zero, Matérn-correlated spatial random effect. The second model (spatial trend model;

log(µ_i_) = log(PT_i_) + β_o_ + β_1_ Trt_i_ + β_2_ lat_i_ + β_3_ lon_i_ + w(s_i_)) augmented the specification with linear fixed effects for latitude (lat_*i*_) and longitude (lon_*i*_), capturing large-scale directional gradients in baseline mortality that the spatial random effect alone might not represent. The third model (linear ATE model; log(µ_i_) = log(PT_i_) + β_o_ + β_1_ Trt_i_ + β_2_ lat_i_ + β_3_ lon_i_ + β_4_(Trt_i_ × lat_i_) + β_5_(Trt_i_ × lon_i_) + w(s_i_)) further added interaction terms, allowing the magnitude of the ATE to vary as a linear function of geographic location. The fourth model 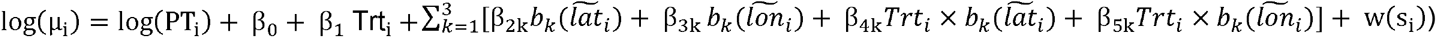 replaced the linear coordinate terms and their treatment interactions with natural cubic spline bases, where *b*⍰(·), *k* = 1, 2, 3, denote the basis functions of a natural cubic spline with three degrees of freedom and 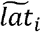 and 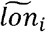 are coordinates centered at the study-area mean. The third model is nested within the fourth, so the comparison tests whether a linear ATE surface is adequate.

Models were compared using the Akaike information criterion (AIC) and, for nested pairs, LRTs; the model with the lowest AIC and a statistically significant improvement over the preceding specification was selected for prediction and all subsequent analyses.

### Spatial predictions and probability of benefit

Estimated mortality surfaces were generated over a 200 x 200 grid spanning Niger’s Dosso region and clipped to its boundary. At each grid point, the selected model was used to predict mortality rates separately under treatment and control. The ATE at a given location was defined as the model-predicted mortality rate under treatment minus the model-predicted mortality rate under control, and the ATE surface was estimated as the pointwise difference between the two predicted mortality surfaces.

To estimate uncertainty in the ATE, we drew 1000 simulated responses from the fitted model, incorporating fixed effects, prediction variance of the Matérn field, and dispersion variance components. At each grid point, the probability of benefit was defined as the proportion of simulated ATEs less than zero, that is, the proportion of simulations in which the treatment was beneficial.

### CSI-level analysis

To assess the implications of the observed spatial heterogeneity in mortality for programmatic surveillance, the modeling framework was repeated on mortality data aggregated at the level of Centre de Santé Intégrés (CSI) catchments. CSIs are primary health centers that serve catchment areas of approximately 25 villages on average, of which only those with a population of 250–2499 were included in AVENIR. Each village was assigned to its nearest CSI by Euclidean distance, and deaths and person-time were aggregated at the CSI level. Each CSI’s location was approximated as the median latitude and longitude across all villages assigned to that CSI. Since treatment group is not defined at the CSI level, this analysis is restricted to assessing spatial aggregation of mortality rates, not treatment effects.

The ICC quantifying the clustering of child mortality within CSIs was estimated from a Poisson generalized linear model: log(µ_i_) = log(PT_i_) + β_₀_ + β_1_ Trt_i_ + u_dept_ + v_CSI|dept_, where *u*_*dept*_ is a random intercept for department and *v*_*CSI* |*dept*_ is a nested random intercept for CSI within department; 95% confidence intervals were obtained from 1000 bootstrap draws. Two Poisson GLMMs were fit: one with an intercept and Matérn spatial random effect, and another with additional linear fixed effects for latitude and longitude. These specifications can be written as log(µ_j_) = log(PT_j_) + β_o_ + w(s_j_) and log(µ_j_) = log(PT_j_) + β_o_ + β_1_lat_j_ + β_2_lon_j_ + w(s_j_), respectively, where j indexes CSIs. AIC and LRT were used to compare the goodness-of-fit of these models. Estimated Matérn spatial ranges from the village- and CSI-level models were compared to characterize the degree to which spatial aggregation attenuates detection of fine-scale structure.

### Spatial heterogeneity in treatment benefit as a function of underlying mortality

To characterize the extent to which spatial heterogeneity in the ATE reflects heterogeneity in underlying mortality, we constructed cumulative gain curves analogous to those used in uplift modeling,^38,39^ where units are ranked by their predicted ATE and cumulative benefit (deaths averted) is plotted against the extent of treatment (**Figure 3**). For each village, doses delivered was defined as the number of children that would have been treated under universal distribution (defined as azithromycin distribution to all children aged 1–59 months in all 1,390 villages at every distribution round over the 24-month follow-up) over all rounds, and deaths averted was the predicted ATE multiplied by person-time at risk; cumulative totals of each were expressed as a proportion of the total under universal treatment. Here, villages were ranked by their predicted ATE and control-group mortality rate under the linear ATE model. Two additional rankings used predicted mortality aggregated to CSI catchments and departments from the same model, with aggregated units ranked by their person-time-weighted predicted control-group mortality. Ranking villages directly on the predicted ATE gives the maximum-attainable curve under this model, and a ranking carrying no information about predicted benefit gives the diagonal.

Heterogeneity captured (H) was defined as the ratio of the area between a predicted mortality-based curve and the diagonal to the area between the maximum-attainable curve and the diagonal, so that H = 1 indicates that predicted benefit is a monotone increasing function of predicted baseline mortality and H = 0 indicates that predicted mortality carries no information about where benefit is concentrated. H is an analogue of the normalized Qini coefficient.^40^

All analyses were conducted in R (version 4.5.2) using the spaMM package^41^ (v 4.6.1). Analysis code is available at [**redacted Zenodo link**].

## Results

### Study population and department-level heterogeneity

The analytic sample comprised 1,390 villages across 8 departments (**Supplementary table 1**): 476 villages assigned to the treatment group and 914 to control (438 villages in the infant azithromycin arm and 476 to the placebo arm). Total follow-up was 276,452 person-years (94,735 treatment group; 181,717 control group). There were 3,635 deaths overall (1,084 treatment; 2,551 control), yielding an overall mortality rate of 13.1 deaths per 1,000 person-years (11.4 treatment; 14.0 control), with an ATE of −2.6 deaths per 1,000 person-years.

Mortality rates (**Figure 2A**; **Supplementary figure 1**) and the ATE varied across departments (**Figure 2D**; **Supplementary figure 2**). Child mortality rates were similar in the infant and placebo arms overall and across departments, supporting pooling them into a single control group (**Supplementary figure 1**). There was significant heterogeneity in mortality rates across departments (LRT χ^2^ = 151.2, *P* < 0.001). The ICC across villages was 0.014 (95% CI: 0.003 to 0.027), indicating modest clustering of mortality rates within departments. Point estimates indicated a reduction in mortality under treatment in all departments except Boboye, where the confidence interval included zero (ATE = 1.4 deaths per 1000 person-years, 95% CI: −1.4 to 4.1; **Supplementary table 1**).

**Figure 2:**
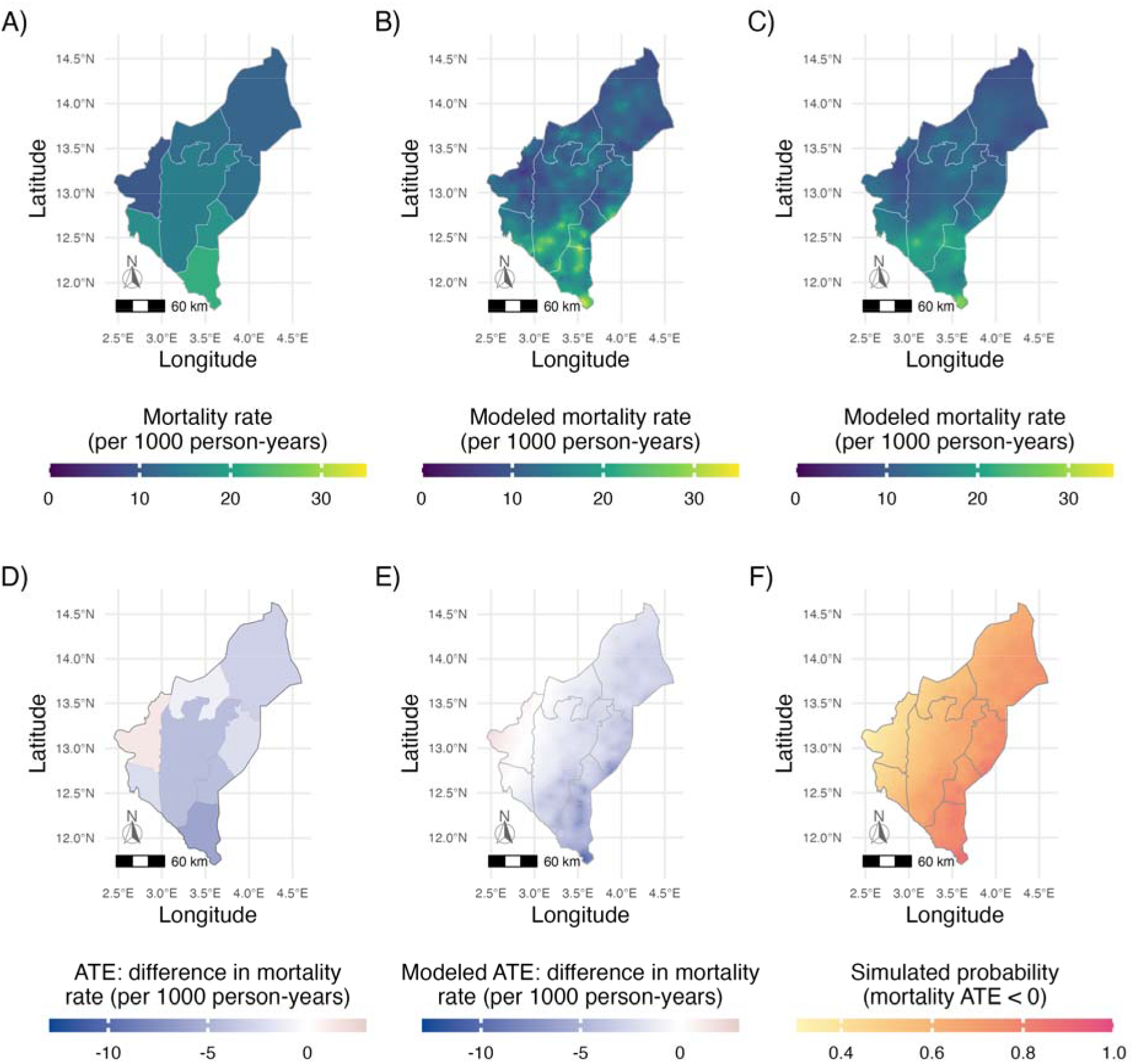
Observed and model-predicted spatial variation in childhood mortality rates, average treatment effects, and posterior probability of treatment being beneficial among children aged 1–59 months in the AVENIR trial in the Dosso region of Niger, 2020-2023. (A) Department-level mortality rates per 1000 person-years averaged over all control villages (randomized to one of infant azithromycin or placebo arms, n = 914 villages). Lighter colors closer to yellow indicate higher mortality rates. Numerical estimates of sample size, rates, and uncertainty and in Supplementary Table 1. (B) Model-predicted mortality rates per 1000 person-years in the control group (n = 914 villages). Predictions were made under the linear ATE model, a Poisson generalized linear mixed model fitted to village-level data that allowed for treatment effects to vary linearly with latitude and longitude and included a Matérn spatial covariance structure to account for spatial autocorrelation in mortality. (C) Model-predicted mortality rates per 1000 person-years using data aggregated to the CSI-level (n = 160 CSIs, 1390 villages). Predictions were made by a Poisson generalized linear mixed model, with a Matern term to account for spatial structure and linear terms for latitude and longitude. (D) Department-level average treatment effects (ATE), defined as the difference between the mortality rates in the treatment and control arms. ATE values range from a beneficial (blue) to unfavorable treatment effect (pink) (E) Model-predicted ATE per 1000 person-years using the same model as panel (B). (**F**) The predicted probability of reduced mortality due to intervention (ATE < 0), based on 1000 simulated draws from the model used in panels B and E. Administrative boundaries used in maps were sourced from the United Nations Office for the Coordination of Humanitarian Affairs (OCHA) *Common Operational Datasets – Administrative Boundaries*, accessed via the Humanitarian Data Exchange on 10 March 2026, and are licensed under CC BY-IGO 3.0 (https://data.humdata.org/dataset/cod-ab-ner). Alt text: Maps of the Dosso region, labelled A to F, showing child mortality rates, average treatment effects, and the probability of treatment benefit, from observed data and model predictions.

### Geospatial models

Village-level spatial GLMMs with latitude and longitude terms outperformed the baseline spatial model by both AIC and LRT (**Supplementary table 2**), consistent with the presence of large-scale spatial trends in mortality not fully captured by residual spatial correlation alone. The model allowing for spatially heterogeneous ATEs using linear treatment interaction terms was preferred over the spatial trends model (LRT χ^2^ = 8.1, *P* = 0.017). The spline ATE model did not improve fit further (LRT χ^2^ = 8.96, *P* = 0.345), indicating that a linear ATE surface was adequate. The linear ATE model was used for all primary analyses.

The estimated Matérn spatial range for the linear ATE model was 11.4 km (*v* = 0.23), essentially identical to the spatial trend model (11.3 km), while the baseline spatial model estimated a substantially longer Matérn range (58.9 km; **Supplementary table 2**), consistent with the Matérn term in the baseline model absorbing spatial variation in mortality rates that the other models can attribute instead to latitude and longitude trends. The spline ATE model had a shorter range (7.6 km) than the linear ATE model, reflecting competition between the spline terms and the Matérn random effect for the same spatial variation.

### Spatial predictions and probability of benefit

Modeled mortality rates showed sub-department heterogeneity, with a pronounced north–south gradient across Dosso with higher rates in the south. This pattern was present across treatment and control surfaces (**Figure 2B**), consistent with spatial variation in baseline mortality rather than an effect of treatment.

The spatial distribution of the predicted ATE diverged from the mortality surface, exhibiting a northwest–southeast gradient (**Figure 2E**). The linear ATE model predicted a localized positive ATE in Boboye, consistent with the observed department-level point estimate. Allowing the ATE to vary with location within the study area permits such localized departures from the overall benefit.

Across 1,000 simulations from the predictive distribution of the linear ATE model, the probability that the ATE was beneficial (ATE < 0) was spatially heterogeneous (**Figure 2F**) and tracked with the predicted mortality reductions under treatment. The probability of benefit was substantially lower in Boboye.

### CSI-level analysis

All included villages were assigned to one of Dosso’s 160 CSIs, with a median of 7 villages per CSI (range: 1–30; **Supplementary figure 3**). After accounting for department-level clustering, the ICC for CSIs within departments was 0.024 (95% CI: 0.016, 0.033), suggesting that sub-department CSI-level variation in mortality contributes consistently to the overall spatial structure. Similar to the village-level models, at the CSI-level, the spatial trends model was preferred over the simple baseline model based on both, AIC and LRT (**Supplementary table 2**).

The Matérn spatial range estimated from the CSI-level spatial trends model was 14.4 km (*v* = 0.4), modestly larger than the estimate from the village-level model with treatment interactions (11.4 km). This inflation in the estimated range is consistent with aggregation smoothing over local spatial variability, effectively increasing the apparent scale of spatial dependence. The CSI-level model predicted mortality surface (**Figure 2C**) preserved the broad north–south gradient detected at the village level, confirming that large-scale patterns are recoverable from administratively aggregated data.

### Spatial heterogeneity in treatment benefit as a function of underlying mortality

Ranking villages by predicted control-group mortality recovered most, but not all, of the spatial heterogeneity of predicted treatment benefit (H = 67.1%; **Figure 3**). Ranking on predicted mortality aggregated to CSIs captured nearly as much heterogeneity as village-level ranking (H = 63.4%), while aggregation to the eight departments captured substantially less (H = 56.1%), consistent with coarser units averaging over the within-unit variation in mortality on which the ranking depends.

**Figure 3:**
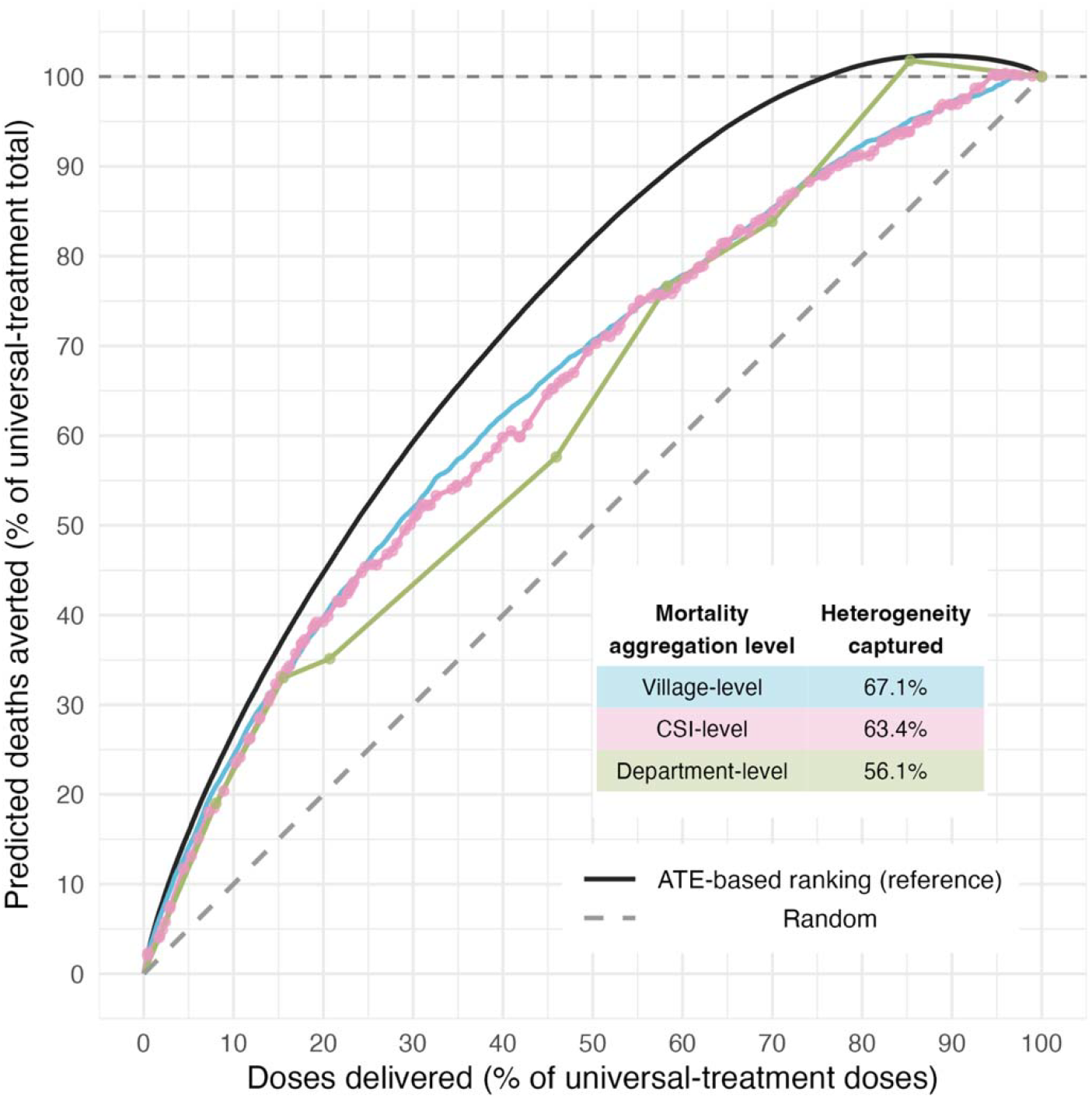
Heterogeneity in predicted treatment benefit captured by heterogeneity in predicted control-group mortality Cumulative gain curves showing predicted deaths averted against doses delivered, expressed relative to universal treatment, defined as azithromycin distribution to all children aged 1–59 months in all 1,390 villages at every round over 24 months. Colored lines rank villages (blue), CSI catchments (pink), and departments (green) by predicted control-group mortality under the village-level linear ATE model. CSI catchments and departments are ranked by their person-time-weighted mean predicted mortality, with points marking unit boundaries and linear interpolation between them. The black line ranks villages by the predicted ATE and represents the maximum attainable concentration of benefit under this model; the gray dashed line (random) represents a ranking that carries no information about predicted benefit. Heterogeneity captured (H), shown in the inset, is the ratio of the area between a mortality-based curve and the random line to the area between the maximum-attainable (ATE-based) curve and the diagonal; H = 100% indicates that predicted benefit is a monotone increasing function of predicted control-group mortality, and H = 0 indicates that predicted mortality carries no information about where benefit is concentrated. Curves may exceed 100% before returning to it at full coverage because predicted benefit is negative in part of the study area. Alt text: Cumulative gain curves of predicted deaths averted against doses delivered, comparing villages, health center catchments, and departments ranked by predicted mortality with a treatment-effect-based reference ranking.

## Discussion

In this secondary analysis of the AVENIR cluster-randomized trial, we found geographic heterogeneity in both child mortality rates and the ATE of azithromycin MDA across Niger’s Dosso region. Mortality rates exhibited a marked north-to-south gradient, with higher rates in the south, while the ATE gradient followed a distinct northwest-to-southeast pattern, with greater treatment benefit in the southeast, which would both be obscured by a single summary effect estimate for the trial. Spatial Poisson GLMMs with Matérn-structured random effects, fit at the village level, provide evidence that this geographic variation in treatment benefit is not adequately captured by aggregate analyses.

The contrast between the north–south mortality gradient and the northwest–southeast ATE gradient is a structural consequence of estimating an absolute effect from a multiplicative model. In the linear ATE model, the treatment contrast is linear on the log scale in latitude and longitude, so the rate ratio follows a smooth, monotone gradient across the study area, varying strongly with longitude and weakly with latitude, and therefore along a roughly east–west axis (**Supplementary figure 4**); allowing this contrast to vary non-linearly with cubic splines did not improve fit. The underlying baseline mortality surface follows a north–south gradient with local departures captured by the Matérn spatial random effect (**Figure 2B**). The predicted ATE, equal to the baseline rate multiplied by (rate ratio − 1), is the product of two surfaces whose gradients are not aligned, and this divergence in orientation generates the oblique ATE surface (**Figure 2E**). Heterogeneity captured (H) assesses the degree to which baseline mortality predicts ATE; the observed intermediate value (H = 67.1%, **Figure 3**) indicates that predicted baseline mortality identifies some, but not all, locations where treatment benefit (ATE) is largest. The differences in geographic trend and spatially local departures identified by the Matérn surface account for the approximately one-third of heterogeneity in ATE not captured by heterogeneity in predicted baseline mortality. Effect measure modification on the absolute scale is arguably the policy-relevant quantity, because, for a fixed number of azithromycin doses distributed, resource allocation could be guided by where the largest absolute reductions in mortality are achievable^42^, and these need not coincide with where baseline mortality is highest or where the relative effect is strongest.

Control-group mortality was markedly lower in Boboye, Falmey, and Loga, which are departments previously treated with azithromycin MDA as part of the MORDOR trial^12,43^ (NCT02047981) compared to departments without recent azithromycin MDA exposure, while treatment-group mortality was similar regardless of prior exposure (**Supplementary figure 5**). Notably, Boboye department had modestly higher mortality in treated than control villages (**Supplementary table 1**). This is consistent with residual protection carried over among control villages that had already received multiple rounds of community-wide azithromycin before AVENIR enrollment, rather than with any reduction in the efficacy of azithromycin MDA. A lower, non-naïve control-group mortality rate in Boboye would also produce a floor effect, whereby populations with lower underlying mortality rate derive smaller marginal benefit from azithromycin MDA; combined with sparse deaths in lower-mortality departments, which may inflate the variance of local effect estimates, a small or null true effect could plausibly be observed as a positive point estimate by chance. The positive ATE in Boboye may therefore reflect a combination of residual protection from prior MDA and statistical imprecision, rather than a true adverse effect of treatment.

CSI-level analysis confirms that programmatic surveillance platforms can recover the broad spatial pattern in the mortality rates despite reducing resolution at local scales. Aggregating outcomes to the CSI level preserved the large-scale spatial structure in the mortality rate, with Matérn range parameters at the CSI level broadly consistent with those estimated at the village level, despite not accounting for treatment effects. However, CSI-level predictions obscured some sub-CSI variation in mortality hotspots— spatially focal areas of elevated mortality risk^44^ —that were detectable at the village level, consistent with the expectation that spatial aggregation smooths local heterogeneity. For programmatic purposes, this tradeoff is likely acceptable: CSIs represent a scalable surveillance infrastructure. Their spatial resolution appears sufficient to guide broad geographic targeting of high-mortality areas. Village-level data and analyses remain necessary to delineate treatment effect heterogeneity.

Geography integrates many ecological, environmental, and sociodemographic characteristics that can jointly modify an intervention’s effect,^35^ which likely accounts for the spatial pattern in treatment benefit reported here. Testing whether specific spatially varying covariates explain this variation would strengthen assessments of generalizability and transportability to other settings.^45^

Some caveats bear on the interpretation of these findings. The analysis draws on data from a single trial conducted in one administrative region of Niger over a 24-month period, and we cannot validate whether the localized mortality patterns identified here are stable over time. The spatial random effects used in our models absorb the fine-scale spatial structure captured by village-level monitoring but cannot decompose it into identifiable causes, so gradients in ATE could be driven by spatially structured effect modifiers such as population density or rainfall. Similarly, heterogeneity captured (H) compares two surfaces predicted from the same fitted model and describes its internal structure rather than out-of-sample performance. Pooling outcomes across the full follow-up period averages over seasonal variation in child mortality and MDA coverage; the estimated mortality and ATE surfaces may thus reflect a mixture of spatial and temporal effects that we did not attempt to disentangle. Matérn-based predictions at the edge of the study area carry greater uncertainty than those in the interior and should be considered when interpreting predictions for border-adjacent regions.

### Conclusions

This analysis revealed spatial heterogeneity in child mortality and the ATE of azithromycin MDA that would be masked by a single trial-wide estimate, with the strongest evidence of benefit concentrated in the southeast of the Dosso region. Aggregating data to CSI-level health catchment areas preserved broad spatial patterns in mortality, supporting a scalable platform for ongoing surveillance. The results demonstrate how spatial models applied to large cluster-randomized trials can identify where treatment effects concentrate and inform study design by indicating the spatial resolution of outcome monitoring.

## Acknowledgements

The AVENIR trial was supported by grants (OPP1210548 and INV-002454) from the Gates Foundation. Pfizer donated azithromycin and placebo. This work was additionally supported in part by the National Institute of Allergy and Infectious Diseases (R01AI175250, R01AI158884 and R01AI166671).

Anthropic Claude Opus 5 was used to review and document analysis code, which was then verified by the authors, and we take full responsibility for the publication.

## Data availability statement

De-identified AVENIR trial data is available through the Open Science Framework (https://osf.io/48q7k/). The village geographic coordinates data required for this analysis are not publicly sharable to protect participant confidentiality. Replication code for all analyses is available through Zenodo (https://doi.org/10.5281/zenodo.23545422). Administrative boundaries used in maps were sourced from the United Nations Office for the Coordination of Humanitarian Affairs (OCHA) *Common Operational Datasets – Administrative Boundaries*, accessed via the Humanitarian Data Exchange on 10 March 2026, and are licensed under CC BY-IGO 3.0 (https://data.humdata.org/dataset/cod-ab-ner).

