## Supplementary Material for "Estimation of fine-scale spatial heterogeneity in treatment effects in a large cluster-randomized trial"

Supplementary tables

1. Mortality rates and ATE by treatment group and department
2. Model specification, fit statistics and estimated parameters

Supplementary figures

1. Department-level child mortality rates by trial treatment arm
2. Department-level average treatment effects (ATEs)
3. **Assignment of villages to health catchment areas**
4. Model-predicted mortality, ATE, and rate ratio surfaces
5. Mortality rates by whether the department was previously included in the MORDOR trial

### SUPPLEMENTARY TABLES

| **Department** | **Treatment group** | | | | **Control group** | | | | **ATE**  **(difference in mortality rates, per 1000py)** |
| --- | --- | --- | --- | --- | --- | --- | --- | --- | --- |
|  | **villages** | **deaths** | **person-years** | **mortality rate  (per 1000py)** | **villages** | **deaths** | **person-years** | **mortality rate  (per 1000py)** |  |
| **Boboye** | 71 | 159 | 14287.2 | 11.1  (95% CI: 8.9, 13.5) | 136 | 276 | 28356.9 | 9.7  (95% CI: 8.3, 11.2) | 1.4  (95% CI: -1.4, 4.1) |
| **Loga** | 47 | 155 | 13274.9 | 11.7  (95% CI: 9.7, 14.0) | 85 | 270 | 21176.9 | 12.7  (95% CI: 10.8, 14.8) | -1.1  (95% CI: -4.0, 1.9) |
| **Tibiri** | 61 | 115 | 10633.5 | 10.8  (95% CI: 8.0, 13.8) | 121 | 283 | 21950.2 | 12.9  (95% CI: 11.1, 14.7) | -2.1  (95% CI: -5.4, 1.5) |
| **Falmey** | 27 | 82 | 5526.1 | 14.8  (95% CI: 10.6, 19.6) | 52 | 169 | 9882.9 | 17.1  (95% CI: 13.9, 20.8) | -2.3  (95% CI: -7.7, 3.4) |
| **Dogondoutchi** | 87 | 123 | 14681.8 | 8.4  (95% CI: 6.7, 10.2) | 154 | 301 | 26370.7 | 11.4  (95% CI: 10.1, 12.8) | -3.0  (95% CI: -5.2, -0.8) |
| **Dioundiou** | 20 | 56 | 4086.3 | 13.7  (95% CI: 9.1, 18.4) | 64 | 304 | 16954.5 | 17.9  (95% CI: 14.7, 21.5) | -4.2  (95% CI: -10.1, 1.5) |
| **Dosso** | 122 | 251 | 23179.2 | 10.8  (95% CI: 9.3, 12.5) | 230 | 666 | 44122.9 | 15.1  (95% CI: 13.4, 16.8) | -4.3  (95% CI: -6.5, -1.9) |
| **Gaya** | 41 | 143 | 9065.9 | 15.8  (95% CI: 12.7, 19.1) | 72 | 282 | 12902.1 | 21.9  (95% CI: 18.0, 25.9) | -6.1  (95% CI: -11.1, -1.0) |
| **Overall** | **476** | **1084** | **94734.7** | **11.4  (95% CI: 10.6, 12.3)** | **914** | **2551** | **181716.9** | **14.0  (95% CI: 13.2, 14.9)** | **-2.6  (95% CI: -3.8, -1.4)** |

**Supplementary Table 1**: Mortality rates in the AVENIR trial by treatment arm and department in the Dosso region of Niger, 2020–2023. Counts of villages, deaths, and person-years at risk are shown separately for the treatment group (child azithromycin arm) and the control group (infant azithromycin and placebo arms pooled). Mortality rates are expressed as deaths per 1,000 person-years among children aged 1–59 months. The Average Treatment Effect (ATE) for each department is defined as the treatment-group mortality rate minus the control-group mortality rate, expressed as deaths per 1,000 person-years. The control group comprises villages randomized to the infant azithromycin arm (n = 438) and the placebo arm (n = 476). 95% confidence intervals are derived from 10000 bootstrap replicates.

| **Aggregation level** | **Model** | **Terms** | **Model Equation** | **AIC** | **LRT** | **rho  (m^-1)** | **range (km)** | **nu** |
| --- | --- | --- | --- | --- | --- | --- | --- | --- |
| Village | Baseline spatial model | Intercept, treatment, Matérn RE | $\log\left( \mu_{i} \right)=\log\left( PT_{i} \right)+\beta_{0}+ \beta_{1}\text{Trt}_{i}+w\left( s_{i} \right)$ | 4997.2  (df: 1088.4) | - | 1.70E-05 | 58.93 | 0.141 |
| Village | Spatial trend model | Baseline spatial model + latitude, longitude | $\log\left( \mu_{i} \right)= \log\left( PT_{i} \right)+ \beta_{0}+ \beta_{1} \text{Trt}_{i}+ \beta_{2} \text{lat}_{i}+ \beta_{3} \text{lon}_{i}+w\left( s_{i} \right)$ | 4988  (df: 1094.1) | χ² = 13.2, p = 0.001 (ref: baseline village model) | 8.87E-05 | 11.27 | 0.226 |
| Village | Linear ATE model | Spatial trend model + treatment interaction with latitude and longitude | $\log\left( \mu_{i} \right)= \log\left( PT_{i} \right)+ \beta_{0}+ \beta_{1} \text{Trt}_{i}+ \beta_{2} \text{lat}_{i}+ \beta_{3} \text{lon}_{i}+ \beta_{4}\left( \text{Trt}_{i}\times\text{lat}_{i} \right)+ \beta_{5}\left( \text{Trt}_{i}\times\text{lon}_{i} \right)+w\left( s_{i} \right)$ | 4983.9  (df: 1098.6) | χ² = 8.1, p = 0.017 (ref: village spatial trend model) | 8.80E-05 | 11.37 | 0.234 |
| Village | Spline ATE model | Linear ATE model with linear terms for latitude and longitude replaced with natural cubic splines (df 3) | $\log\left( \mu_{i} \right)= \log\left( PT_{i} \right)+ \beta_{0}+ \beta_{1} \text{Trt}_{i}+ \sum_{k=1}^{3} \beta_{2^{k}}f_{k}\left( \text{lat}_{i} \right)+ \sum_{k=1}^{3} \beta_{3^{k}}g_{k}\left( \text{lon}_{i} \right)+ \sum_{k=1}^{3} \beta_{4^{k}}\left( \text{Trt}_{i}\times f_{k}\left( \text{lat}_{i} \right) \right)+ \sum_{k=1}^{3} \beta_{5^{k}}\left( \text{Trt}_{i}\times g_{k}\left( \text{lon}_{i} \right) \right)+w\left( s_{i} \right)$ | 4990.9 (df: 1097.0) | χ² = 9.0, p = 0.345 (ref: linear ATE model) | 1.32E-04 | 7.59 | 0.295 |
| CSI | Baseline spatial model | Intercept, Matérn RE | $\log\left( \mu_{j} \right)= \log\left( PT_{j} \right)+ \beta_{0}+w\left( s_{j} \right)$ | 1052.32  (df: 82.3) | - | 6.01E-06 | 166.39 | 0.203 |
| CSI | Spatial trend model | Baseline spatial model + latitude, longitude | $\log\left( \mu_{j} \right)= \log\left( PT_{j} \right)+ \beta_{0}+ \beta_{2} \text{lat}_{j}+ \beta_{3} \text{lon}_{j}+w\left( s_{j} \right)$ | 1045.79 (df: 83.4) | χ² = 10.53, p = 0.005 (ref: baseline CSI model) | 6.96E-05 | 14.37 | 0.396 |

**Supplementary Table 2**: Model fit statistics and estimated Matérn spatial parameters for Poisson generalized linear mixed models to estimate the Average Treatment Effect (ATE) defined as the incidence rate difference, fit at the village and CSI levels. For village-level models, four specifications are reported: a baseline spatial model, a spatial trend model adding linear fixed effects for latitude and longitude, a linear ATE model with treatment × latitude and treatment × longitude interaction terms, and a spline ATE model, in which the linear latitude and longitude terms and their treatment interactions are replaced by natural cubic spline bases with 3 degrees of freedom. For CSI-level models, two specifications are reported: a baseline spatial model with an intercept and Matérn spatial random effect, and a spatial trend model adding linear fixed effects for latitude and longitude. Treatment indicators were not included in CSI-level models because randomization occurred at the village level and treatment assignment is not defined at the CSI level. In the model equations, $i$ indexes villages and $j$ indexes CSIs; the observed death count is modeled as Poisson-distributed with expectation $\mu$, with $PT$, the person-years at risk, entered as an offset on the log scale; $Trt$ is the binary treatment group indicator; $lat$ and $lon$ are geographic coordinates; f(·) and g(·) denote the natural cubic spline bases of latitude and longitude, respectively, with k indexing the three basis functions of each, and $w\left( s \right)$ is a mean-zero, Matérn-correlated spatial random effect evaluated at each location. For each model, the table reports the Akaike information criterion (AIC), effective degrees of freedom (df), likelihood ratio test (LRT) statistic and p-value relative to the preceding nested model, the estimated Matérn range parameter (km), and the Matérn smoothness parameter (nu). All models were fit by maximum likelihood using the spaMM package (v4.6.1) in R (v 4.5.2), with the log of person-years as an offset and no nugget term.

**SUPPLEMENTARY FIGURES**

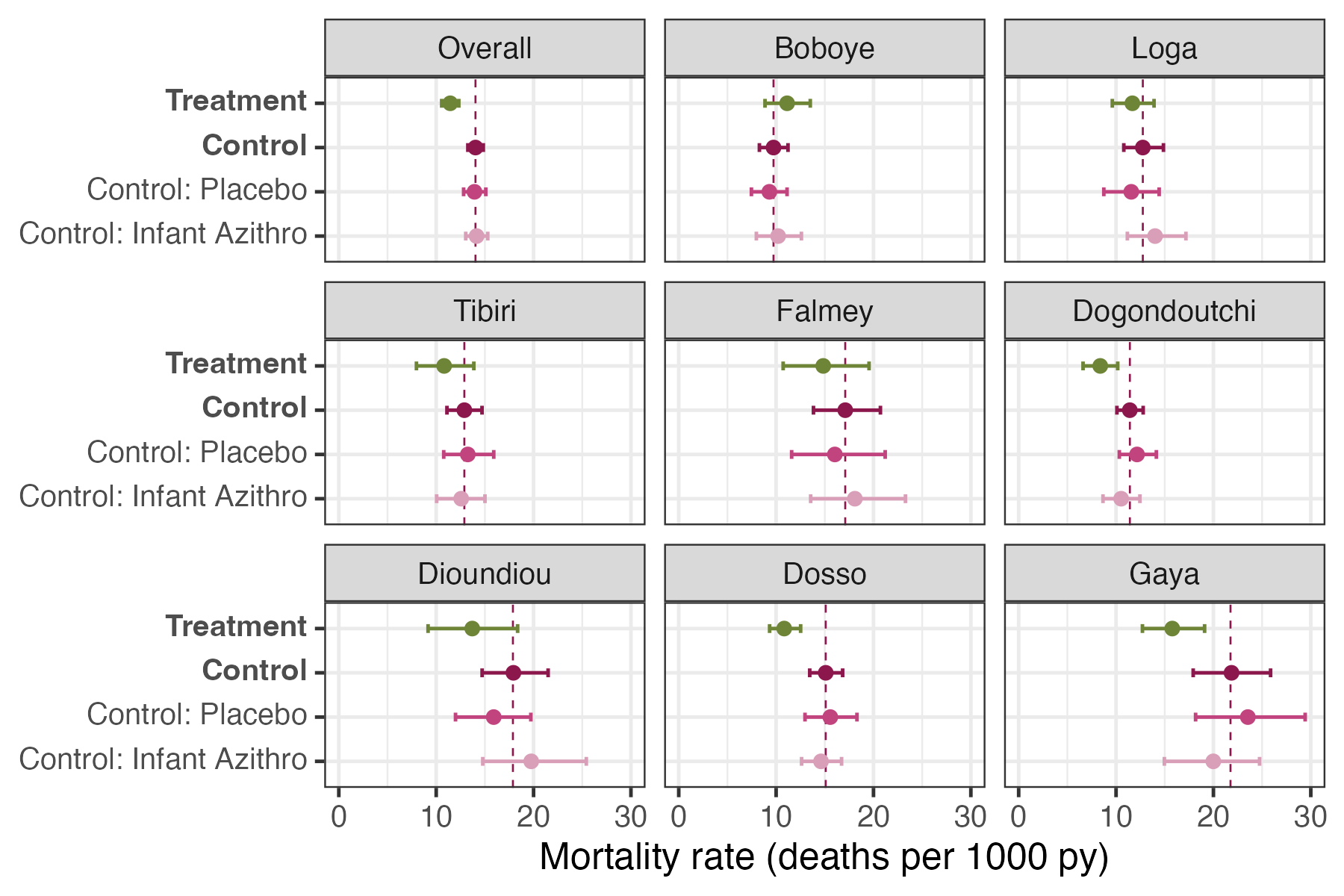

**Supplementary Figure 1**: Department-level child mortality rates in the AVENIR trial in Dosso region, Niger, 2020–2023. Mortality rates are shown separately for treatment (child azithromycin) and control (infant azithromycin and placebo pooled) villages, expressed as deaths per 1,000 person-years among children aged 1–59 months. Mortality rates in the control group are also shown disaggregated as the infant azithromycin and placebo trial arms, which were pooled on the basis that the infant arm did not achieve a statistically significant reduction in child mortality in the primary trial analysis. Points represent department-level rate estimates; error bars indicate 95% confidence intervals derived from 10000 bootstrap replicates. Vertical dashed lines represent the pooled control group mortality rate. A likelihood ratio test based on Poisson models indicated statistically significant heterogeneity in mortality rates across departments (χ² = 151.2, p < 0.001).

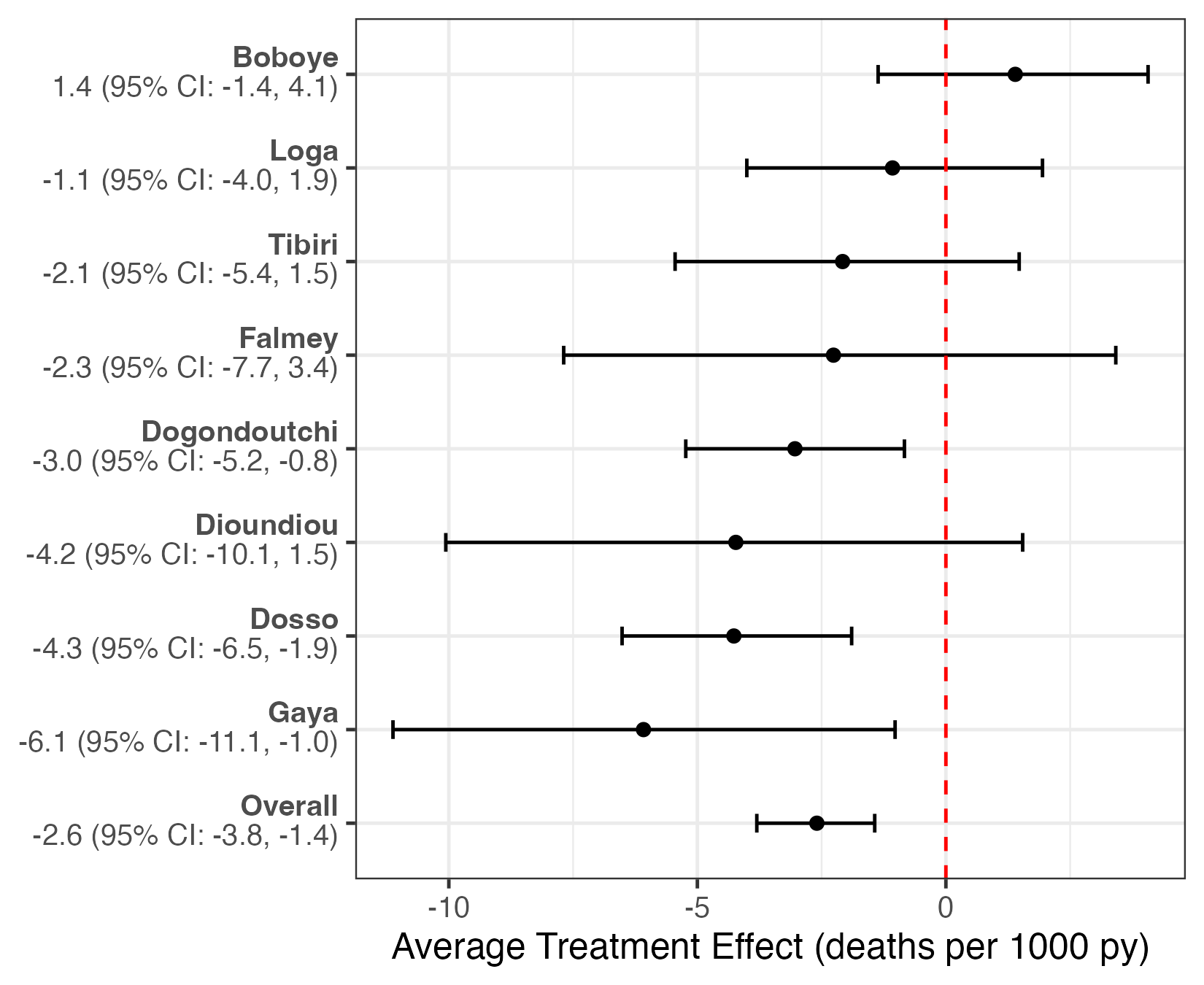

**Supplementary Figure 2**: Department-level average treatment effects (ATEs) of azithromycin MDA in the AVENIR trial, Dosso region, Niger, 2020–2023. The ATE for each department is defined as the treatment-group mortality rate minus the control-group mortality rate, expressed as deaths per 1,000 person-years among children aged 1–59 months. The control group in this analysis comprises villages randomized to the infant azithromycin arm (n = 438) and the placebo arm (n = 476). Error bars indicate 95% confidence intervals estimated by bootstrapping. Negative values indicate a beneficial effect of azithromycin. All departments except Boboye showed a reduction in mortality under treatment. Departments are ordered along the x-axis from most to least beneficial ATE. A likelihood ratio test based on Poisson models indicated statistically significant heterogeneity in ATEs across departments (χ² = 18.4, p = 0.01).

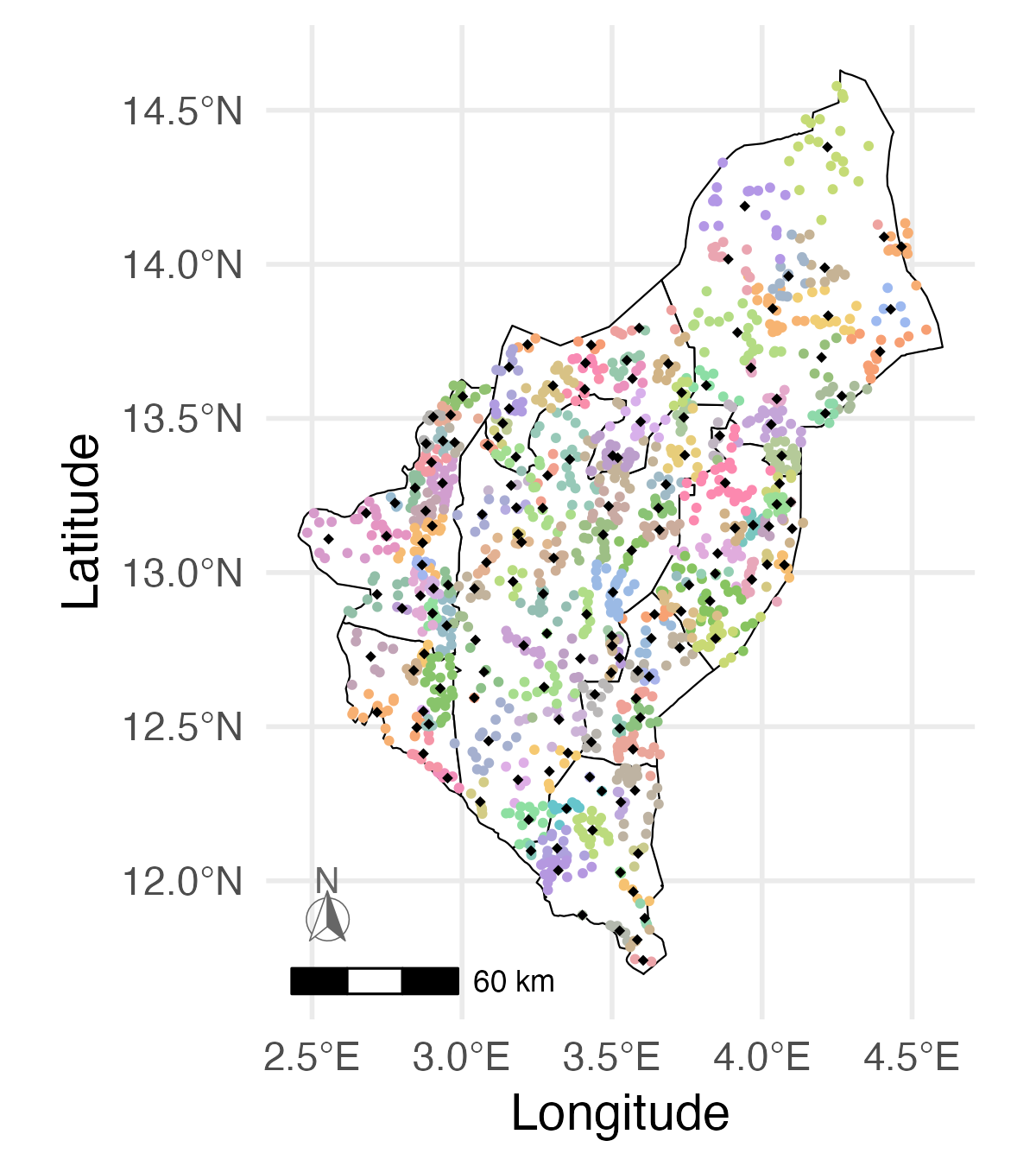

**Supplementary Figure 3**: **Assignment of villages to health catchment areas (Centres de Santé Intégrés, CSIs) in the AVENIR trial, Dosso region, Niger, 2020–2023.** Villages included in the analytic sample are colored by the CSI to which each was assigned; black diamonds indicate the approximate location of each CSI, calculated as the centroid of its assigned villages. Villages were assigned to their nearest CSI by Euclidean distance, and deaths and person-years were aggregated within each resulting catchment area for the CSI-level analysis. Adjacent villages sharing the same color indicate assignment to the same CSI, illustrating that most catchment assignments are spatially contiguous.

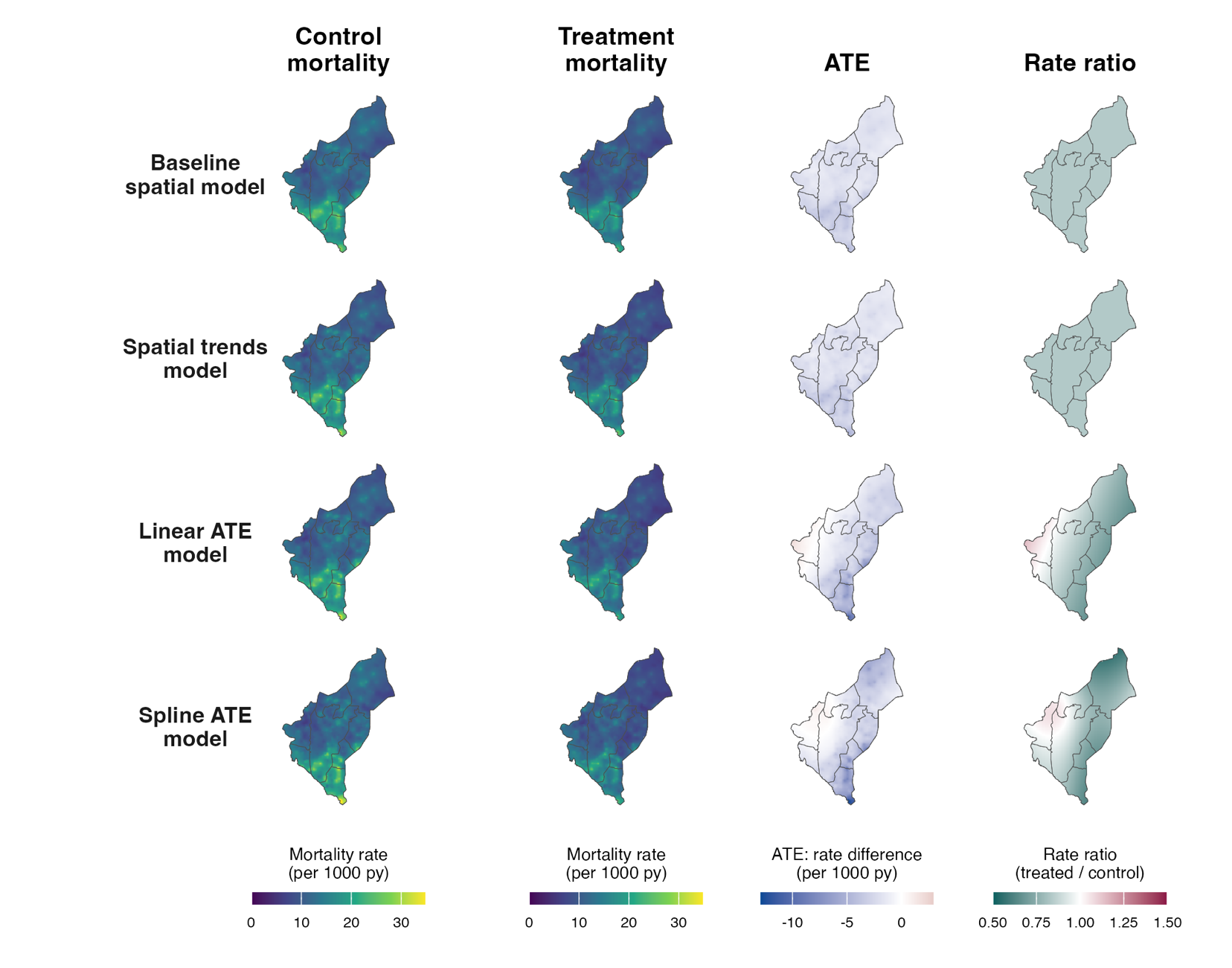

**Supplementary figure 4:** Model-predicted mortality, average treatment effect (ATE), and rate ratio surfaces under four spatial model specifications, AVENIR trial, Dosso region, Niger, 2020–2023.

Rows correspond to the four Poisson generalized linear mixed models fit at the village level: the baseline spatial model, the spatial trends model, the linear ATE model (primary model used for Figure 2B, 2E, and 2F), and the spline ATE model. Model specifications and fit statistics are given in Supplementary table 2.

Columns show, at each grid point: predicted child mortality rate under control; predicted mortality rate under treatment (both per 1,000 person-years among children aged 1–59 months); the ATE, defined as the predicted mortality rate in the treatment group minus the predicted mortality rate in the control group, where negative values (blue) indicate a beneficial effect of azithromycin; and the rate ratio, defined as the predicted mortality rate in the treatment group divided by the mortality rate in the control group, where values below 1.0 (green) indicate a beneficial effect and values above 1.0 (red) indicate higher predicted mortality under treatment. Predictions were generated on a continuous 200 × 200 grid spanning the bounding box of the Dosso region and clipped to its administrative boundary; grey lines indicate department boundaries. Color scales are shared across rows within each column.

The predicted mortality surfaces are visually indistinguishable across the four model specifications: all four recover the same north–south gradient with similar local features, in both arms. This is expected, since baseline mortality is dominated by the offset, intercept, and Matérn random field, which are common to all four models, and the treatment contrast is small relative to the baseline rate.

In the baseline spatial and spatial trends models, treatment enters only as a main effect, so the rate ratio is constant across the study area by construction and all spatial variation in the ATE surface is inherited from variation in the baseline mortality surface. Allowing the log rate ratio to vary with location, in either the linear or spline specification, produces a smooth gradient along a roughly east–west axis, with relative benefit largest in the east and attenuating westward, and a localized region of predicted excess mortality under treatment at the western edge of the study area corresponding to Boboye department. The spline specification recovers a broadly similar gradient with modest additional curvature, indicating that the linear ATE specification captures the dominant spatial structure in the treatment contrast.

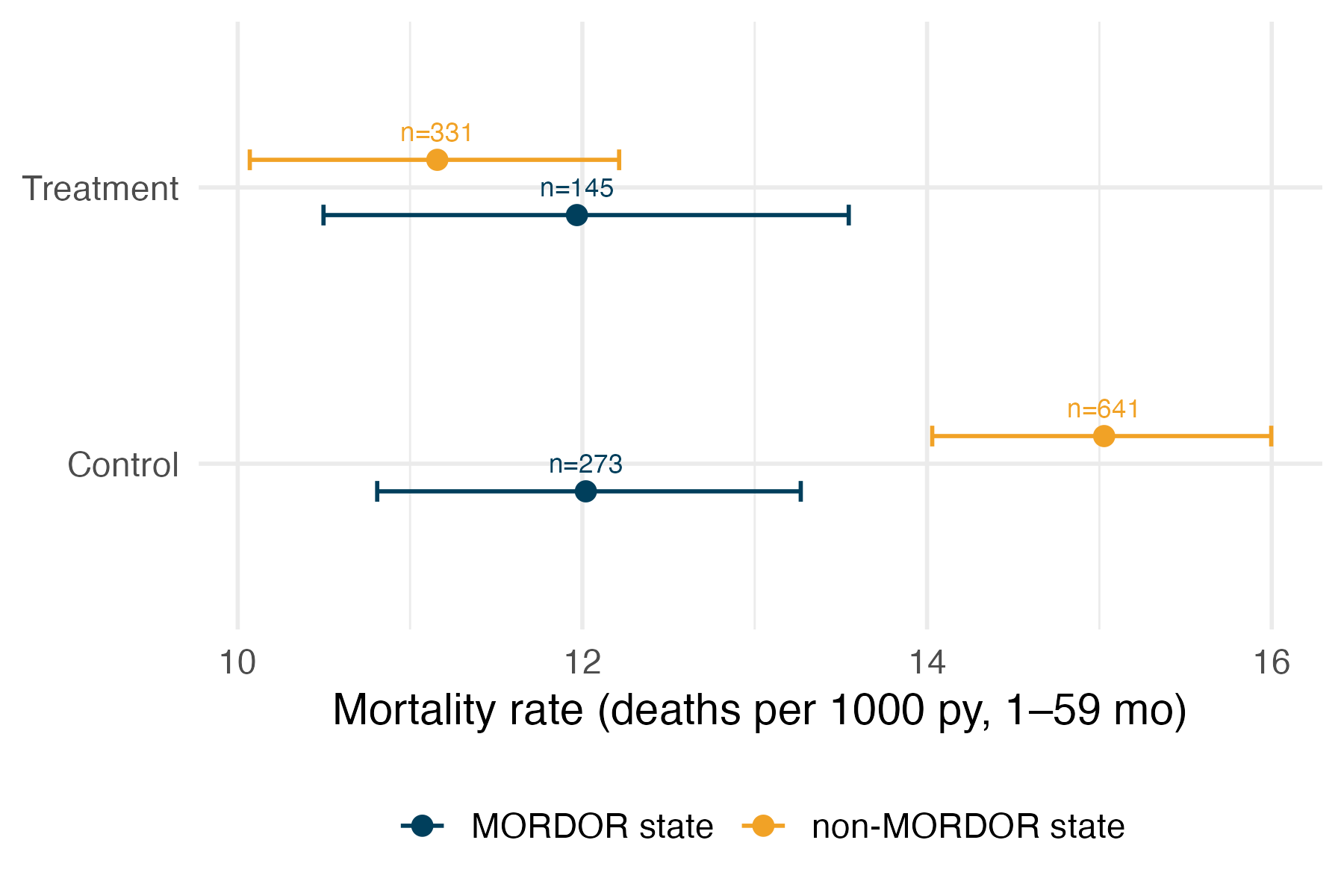

**Supplementary Figure 5:** Child mortality rates by treatment group in AVENIR departments previously included in the MORDOR trial (Boboye, Falmey, and Loga) versus departments not previously included in MORDOR. Points represent mortality rates per 1,000 person-years among children aged 1–59 months in the treatment (child azithromycin arm) and control (infant azithromycin and placebo arms) groups; error bars indicate 95% confidence intervals derived from 1000 bootstrap replicates. Mortality in the treatment group was similar regardless of prior MORDOR exposure, but control-group mortality was markedly lower in previously MORDOR-treated departments than in non-MORDOR departments, consistent with residual protection carried over from prior community-wide azithromycin distribution among control villages.
